# Annotation-free multi-organ anomaly detection in abdominal CT using free-text radiology reports: A multi-center retrospective study

**DOI:** 10.1101/2024.06.10.24308633

**Authors:** Junya Sato, Kento Sugimoto, Yuki Suzuki, Tomohiro Wataya, Kosuke Kita, Daiki Nishigaki, Miyuki Tomiyama, Yu Hiraoka, Masatoshi Hori, Toshihiro Takeda, Shoji Kido, Noriyuki Tomiyama

## Abstract

**Background:** Artificial intelligence (AI) systems designed to detect abnormalities in abdominal computed tomography (CT) could reduce radiologists’ workload and improve diagnostic processes. However, development of such models has been hampered by the shortage of large expert-annotated datasets. Here, we used information from free-text radiology reports, rather than manual annotations, to develop a deep-learning-based pipeline for comprehensive detection of abdominal CT abnormalities.

**Methods:** In this multicenter retrospective study, we developed a deep-learning-based pipeline to detect abnormalities in the liver, gallbladder, pancreas, spleen, and kidneys. Abdominal CT exams and related free-text reports obtained during routine clinical practice collected from three institutions were used for training and internal testing, while data collected from six institutions were used for external testing. A multi-organ segmentation model and an information extraction schema were used to extract specific organ images and disease information, CT images and radiology reports, respectively, which were used to train a multiple-instance learning model for anomaly detection. Its performance was evaluated using the area under the receiver operating characteristic curve (AUC), accuracy, sensitivity, specificity, and F1 score against radiologists’ ground-truth labels.

**Findings:** We trained the model for each organ on images selected from 66,684 exams (39,255 patients) and tested it on 300 (295 patients) and 600 (596 patients) exams for internal and external validation, respectively. In the external test cohort, the overall AUC for detecting organ abnormalities was 0·886. Whereas models trained on human-annotated labels performed better with the same number of exams, those trained on larger datasets with labels auto-extracted via the information extraction schema significantly outperformed human-annotated label-derived models.

**Interpretation:** Using disease information from routine clinical free-text radiology reports allows development of accurate anomaly detection models without requiring manual annotations. This approach is applicable to various anatomical sites and could streamline diagnostic processes.

**Funding:** Japan Science and Technology Agency.

## Introduction

Computed tomography (CT) is an essential diagnostic tool in various clinical settings. CT use has increased globally, with the OECD reporting an average increase exceeding 60% in 2021.^1^ Thorough search of CT images for abnormalities is a critical routine practice for radiologists; thus, the increase in scans implies increased radiologists’ workload.^2,3^ Moreover, increased workload correlates with inter-reader interpretive discrepancies.^4^ Accordingly, new technological approaches are needed to streamline diagnostic processes and enhance their accuracy.

Many studies have demonstrated the utility of artificial intelligence (AI) to improve radiologists’ workflow^5^ and diagnostic accuracy.^6^ However, developing accurate AI diagnostic support systems with deep-learning requires training with a large labeled datasets.^7^ Annotating medical images is labor-intensive and time-consuming, and also requires medical expertise to be effective.^8^ Furthermore, the scarcity of medical data combined with patient privacy issues limits public dataset availability. Accordingly, innovative approaches are needed to overcome the lack of annotated data to advance medical AI.

The annotation burden can be alleviated by reusing information from medical records obtained during routine clinical practice.^9^ After natural language processing to extract pertinent disease information, free-text radiology reports of chest X-ray or head CT images have been used as effective labels, reducing the need for manual annotation.^10–13^ However, applying this method to abdominal CT images is difficult, because of their complexity. Abdominal CT, which captures extensive body areas and multiple organs in three-dimensional (3D) images, comprises tens of millions of voxels and varies substantially in individual characteristics and imaging conditions. This complexity necessitates a refined approach to identify each organ accurately, and extract and link disease information from free-text reports. Additionally, the scarcity of large datasets that pair abdominal CT images with detailed reports hampers AI research and development of effective diagnostic support systems.

Here, we propose a fully end-to-end pipeline for detecting abnormalities in five organs (the liver, gallbladder, kidney, spleen, and pancreas). We first extracted organ-specific regions from CT images using a multiorgan segmentation model, which enables focused predictions of specific organs. We then employed an information extraction schema to derive disease information for each organ from radiology reports and used this information as a training label, thereby eliminating the need for additional manual annotations.

## Methods

### Study design and participants

Our pipeline criteria followed the checklist for Artificial Intelligence in Medical Imaging criteria.^14^ In this retrospective study, data were collected from nine institutions—The University of Tokyo, Keio University, Okayama University, Ehime University, Juntendo University, Kyoto University, Kyushu University, Osaka University, and Tokushima University—using the Japan Medical Imaging Database (J-MID). This multicenter study was approved by the ethics committees of each institution, and anonymized data were exchanged among the institutions under a data-sharing agreement. The need for obtaining written informed consent was waived because of the retrospective data acquisition from the J-MID.

### Inclusion criteria and data split

Patient enrollment is summarized in Figure 1. We used abdominal CT images collected from the nine institutions during routine clinical practice, from July 1, 2020, to February 27, 2023. Images with a large number of slices (>300) or a small number of slices (<40) were excluded to improve computational efficiency. Axial abdominal exams were selected according to the protocol names and outputs from the information extraction schema. These images were divided into internal training, internal test, and external test cohorts.

**Figure 1.**
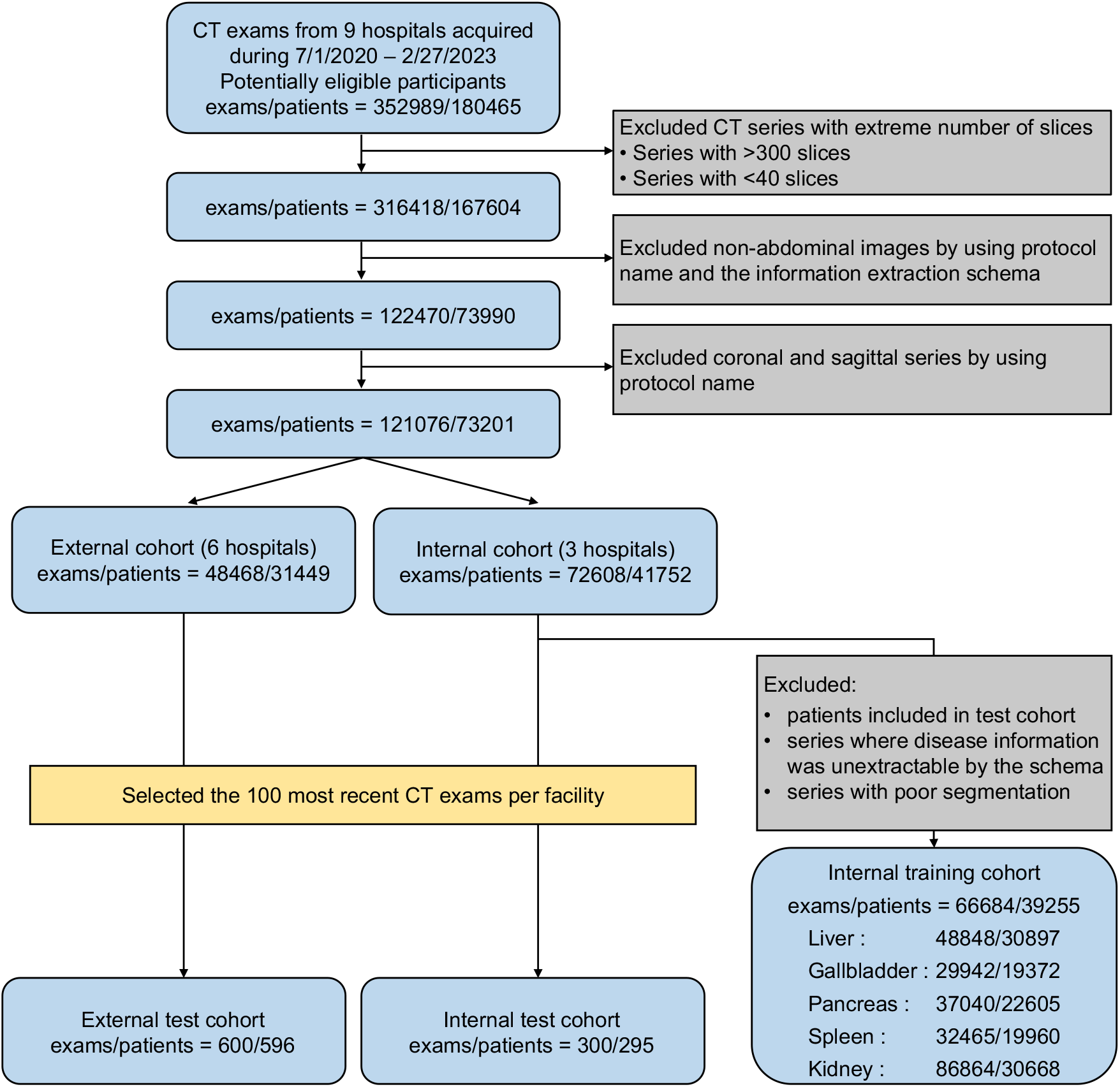
Flowchart of patient enrollment If some exams contained multiple series, such as non-contrast and contrast-enhanced images, all series that met the inclusion criteria were used. The kidney images were counted separately for the left and right sides.

Among them, images from three institutions were designated as the internal test cohort and those from the remaining six as the external test cohort. For both internal and external test cohorts, the 100 most recent exams from each institution were selected. Data from patients assigned to the internal test cohort were excluded from the internal training cohort. Additionally, images with poor segmentation or those from which disease information could not be extracted using a structured protocol were excluded from the internal test cohort.

### Anomaly detection pipeline overview

We used a three-stage approach: labeled dataset extraction, model training, and inference (Figure 2). We curated a dataset comprising abdominal CT images accompanied by radiology reports and associated patient information from the J-MID database. We then trained a deep-learning-based pipeline to detect abnormal findings across five abdominal organs. During training, 3D CT images were input into a multi-organ segmentation model that extracted specific organ information from the entire abdominal image (Figure 2A). The accompanying radiology reports were processed using an information extraction schema to determine the presence or absence of diseases in these organs (Figure 2B). We then trained a model to detect diseases in each organ using organ-cropped images as input and the presence of a disease as a training label (Figure 2C). During inference, combining the segmentation and anomaly detection models enabled processing of whole abdominal images to calculate abnormality scores for each organ, which could assist physicians in their diagnoses (Figure 2D). These steps are detailed below.

**Figure 2.**
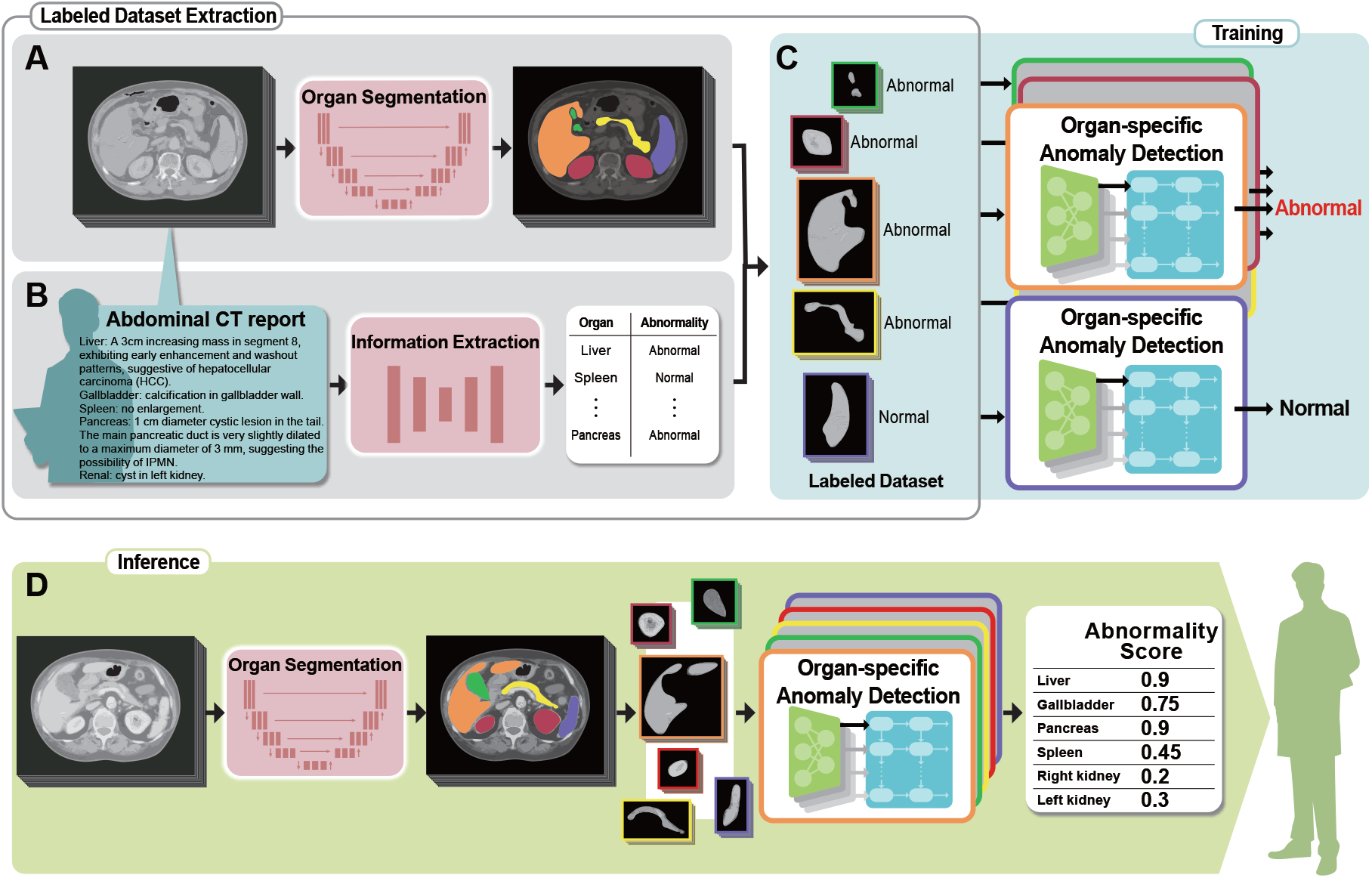
Our anomaly detection pipeline using free-text radiology reports. (A) In the training process, abdominal computed tomography (CT) images are first input into a multi-organ segmentation model. (B) Radiology reports corresponding to the CT images are checked for the presence of disease in each organ using an information-extraction schema. (C) The extracted disease information is used as supervised labels for training of the model with images of the organs. (D) The trained model computes anomaly scores for individual organs from images, offering diagnostic assistance.

### Multi-organ segmentation model

Organ segmentation is effective for efficiently training anomaly detection models by extracting organs from large images, such as abdominal images. This approach has performed well in previous AI tasks for estimating cervical spine fractures.^15^ We adopted a custom 3D full-resolution variant of nnUNet,^16^ which enables accurate segmentation with big patch and batch sizes.^17^ Training was performed using a batch size of 16 and a patch size of 288 × 288 × 64 pixels (width, height, and depth), with all other settings following the original nnUNet study parameters. The model was trained to segment 13 anatomical structures: the liver, gallbladder, pancreas, spleen, left and right kidneys, esophagus, stomach, duodenum, aorta, left and right adrenal glands, bladder, and prostate/uterus. The model was trained and evaluated with 431 images from 2 sources: 300 images from AMOS,^18^ a large-scale and diverse clinical dataset for abdominal organ segmentation (200 for training and 100 for testing), and 131 images from the Computational Anatomy Project dataset^19^ (117 for training and 14 for testing).

### Information extraction schema

A large amount of expert-annotated training data is required to develop deep-learning models. To reduce this requirement, we used an information extraction schema to reuse existing medical data. We hypothesized that the information on the presence of abnormal findings obtained from this schema could serve as a surrogate training label. As demonstrated in our previous study using deep-learning-based named-entity recognition,^20^ the information extraction schema includes not only clinical findings, but also modifiers that detail the involved organ as well as their sizes and characteristics. Moreover, each clinical finding was assigned a certainty score to indicate the level of confidence in its presence, which ranged from 0 (definite absence) to 4 (definite presence). Absence of lesions (score 0) was categorized as “no finding,” whereas any sign of potential lesions (scores 1–4) was classified as an “abnormal finding.” The algorithm was described in detail in our previous paper.^21^ Subsequently, patients with abnormal findings were categorized. Radiologists defined several disease categories for each organ and determined the most appropriate category for each extracted disease word. The correspondence table between words indicating abnormal findings and their respective categories is presented in the Data Sharing section.

### Training protocol and preprocessing of anomaly detection model

Anomaly detection models were trained to identify organ abnormalities, with input of segmented 3D organ images, and output of disease categories. Such training was conducted for each organ through multiclass multi-label learning with labels generated by the information extraction schema (Supplementary Figure 1). 3D images were processed using multiple-instance learning,^22^ input into a 2D encoder at set slice intervals, and information across slices was integrated to obtain the final class output. Organ-segmented 3D images, where non-organ voxels were filled with background values (−1000), were resized to 256 × 256 × 64 pixels in the left–right, antero–posterior, and cranio–caudal axes. From these 3D images, sets of five adjacent slices were extracted to form an image with five channels, which were then inputted into a 2D convolutional neural network (CNN) model in two-slice steps (0^th^, 2^nd^, 4^th^, …). The outputs were inputted to a Long Short-Term Memory network to share information between slices, with the final class output generated through fully connected layers and global average pooling. As 2D CNN encoder, ConvNeXt v2^23^ was used. Window-level and window-width were set to 100 and 300, respectively, and normalized to a range between -1 and 1 before model input.

Training was performed over 12 epochs, using 5-fold cross-validation to select the model with the highest area under the receiver operating characteristic (ROC) curve (AUC) on the validation dataset as the final model for each fold. The final prediction of the test cohort was based on the average of the five models. The threshold for anomaly prediction was determined using the median of the thresholds that yielded the highest F1 scores across the validation dataset for each fold. Cross-entropy loss and the AdamW optimizer were employed with a learning rate of 0·00023 and a cosine-annealing learning-rate scheduler. The anomaly detection model was trained using a computing node with two CPUs and eight NVIDIA A100 graphics cards.

### Ground-truth annotation

Each exam in our dataset included a free-text report written by a board-certified radiologist. First, to evaluate the accuracy of our information extraction schema on free-text radiology reports, abnormalities were extracted through review by a radiologist. Abnormalities were defined as findings previously documented as explicitly present. Indirect signs suggestive of abnormalities around the organ were not included. For the test cohort of 900 cases, 2 radiology residents reviewed the reports for each organ (considering the left and right kidneys separately) and listed the abnormalities. A third board-certified radiologist resolved any disagreements.

To evaluate the performance of our anomaly detection model, CT images were reviewed to create ground-truth labels. Although the exams already included reports from board-certified radiologists, another radiology resident double-checked them for diagnostic accuracy. In cases of discrepancies or judgment difficulties between the information extraction schema and the resident’s review, a board-certified radiologist reviewed the images for a final decision. The ground-truth established here was based on abnormalities identified by radiologists from CT images, rather than on pathologically confirmed abnormalities. Images were annotated using the SYNAPSE SAI Viewer (FUJIFILM Corporation, Minato, Japan).

### Statistical analysis

The segmentation model’s performance was evaluated using the Dice similarity coefficient (DSC) and normalized surface Dice (NSD) score, as employed in AMOS. The AI system performance was assessed using the following metrics: AUC, accuracy, sensitivity, specificity, and F1 score. These metrics were calculated using Python (v3.9.12; https://www.python.org/downloads/release/python-3912/), NumPy (v1.23.2; https://numpy.org/), and scikit-learn (v1.0·2; https://scikit-learn.org/stable/whats_new/v1.0·html) packages.

Confidence intervals for performance metrics were calculated as the 2.5th and 97.5th percentile of 1000 bootstraps, resampled with replacements from the test cohorts. The agreement between our information extraction schema and the ground-truth labels for creating the training labels was calculated using Cohen’s kappa coefficient. To evaluate the impact of dataset size on accuracy, Delong’s test was performed. The code and abnormal class information used for the implementation of our anomaly detection pipeline are available at (https://github.com/jun-sato/sato_j-mid_ad).

## Results

### Training and evaluation of multi-organ segmentation models

We trained and evaluated a 3D-multi-organ segmentation model using 317 CT images from the AMOS and Computational Anatomy Project datasets for training and 114 images for validation. Boxplots for the DSC and NSD scores across the six organs for segmentation of the test data are presented in Figure 3a and 3b, respectively. For all organs, the median DSC exceeded 90%, with the liver showing the highest DSC (0·981 [25^th^–75^th^ percentile: 0·974–0·986]) and the pancreas showing the lowest DSC (0·911 [0·882–0·934]). The spleen had the highest NSD median value (0·934 [0·902–0·965]), whereas the pancreas had the lowest value (0·770 [0·706–0·831]). Representative examples of test data segmentation are shown in Figure 3c. Detailed information on the segmentation results is provided in Supplementary Table 1.

**Figure 3.**
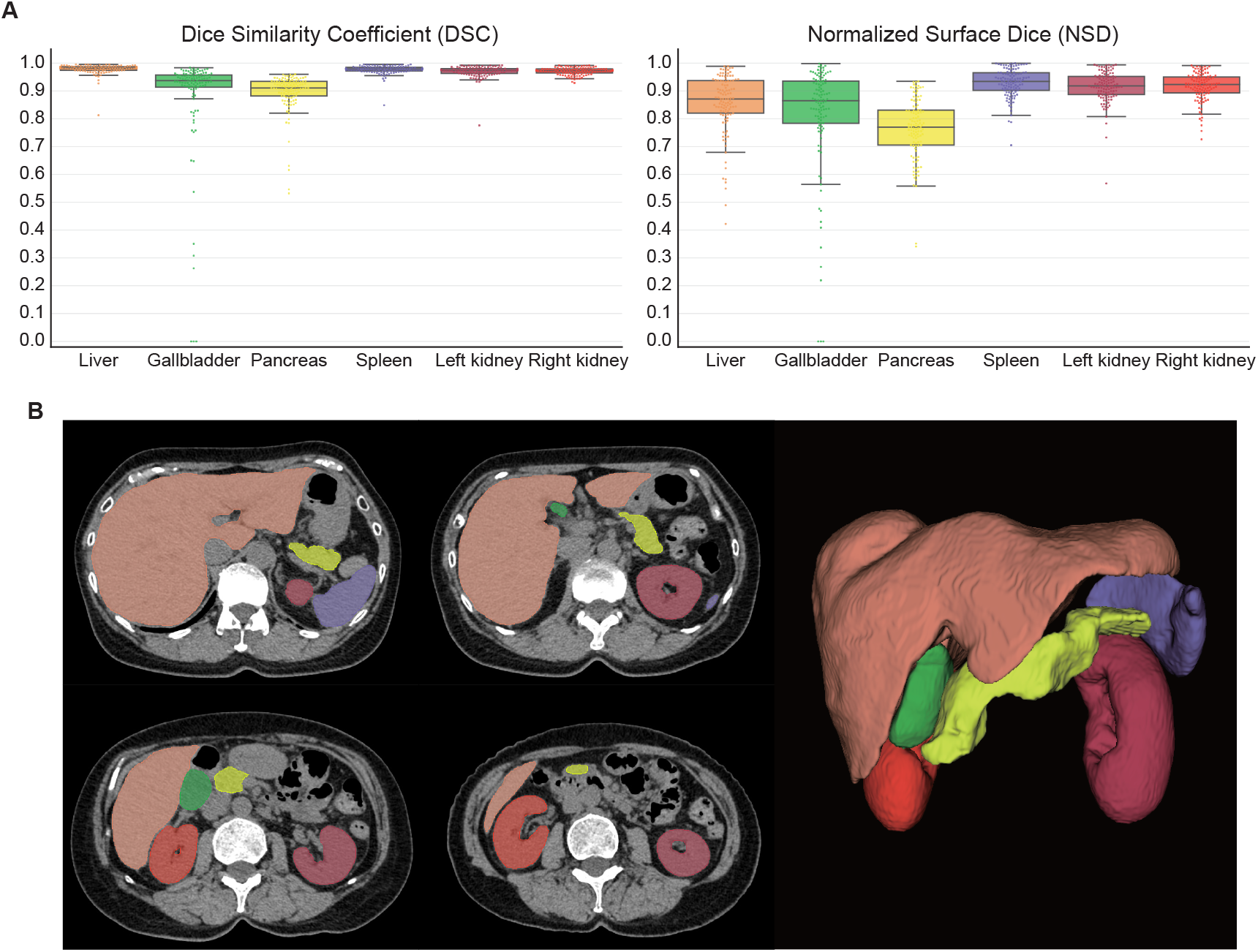
Multi-organ segmentation performance plots and representative examples. (A) Box plots and swarm plots of Dice similarity coefficient (DSC) and normalized surface Dice (NSD) scores in our organ segmentation model. Box plots are defined as follows: the box’s lower and upper bounds are represented by the first and third quartiles of the dataset, respectively, with a median line positioned at the center. Whiskers stretch from the box up to a maximum of 1.5 times the interquartile range, and down to the minimum and up to the maximum data points lying within this range. (B) Axial computed tomography images and three-dimensional scans with mapped model predictions.

### Performance of the structured model on free-text radiology reports

We applied our information extraction schema to all free-text radiology reports accompanying included CT exams (Figure 4a). Within the 900 exams used for our internal and external test cohorts, we examined the extent to which organ-specific abnormal findings, extracted using the information extraction schema, matched the ground-truth verified by at least two radiologists. We assessed the performance of the language model in binary classification by identifying the presence or absence of abnormal findings. The accuracy, sensitivity, specificity, F1 score, and confusion matrices of the information extraction schema are presented in Figure 4b and 4c. The F1 score ranged between 93.1 and 98.5 across organs. The agreement with radiologist annotations was high (Cohen’s kappa coefficients: 0·844–0·942).

**Figure 4.**
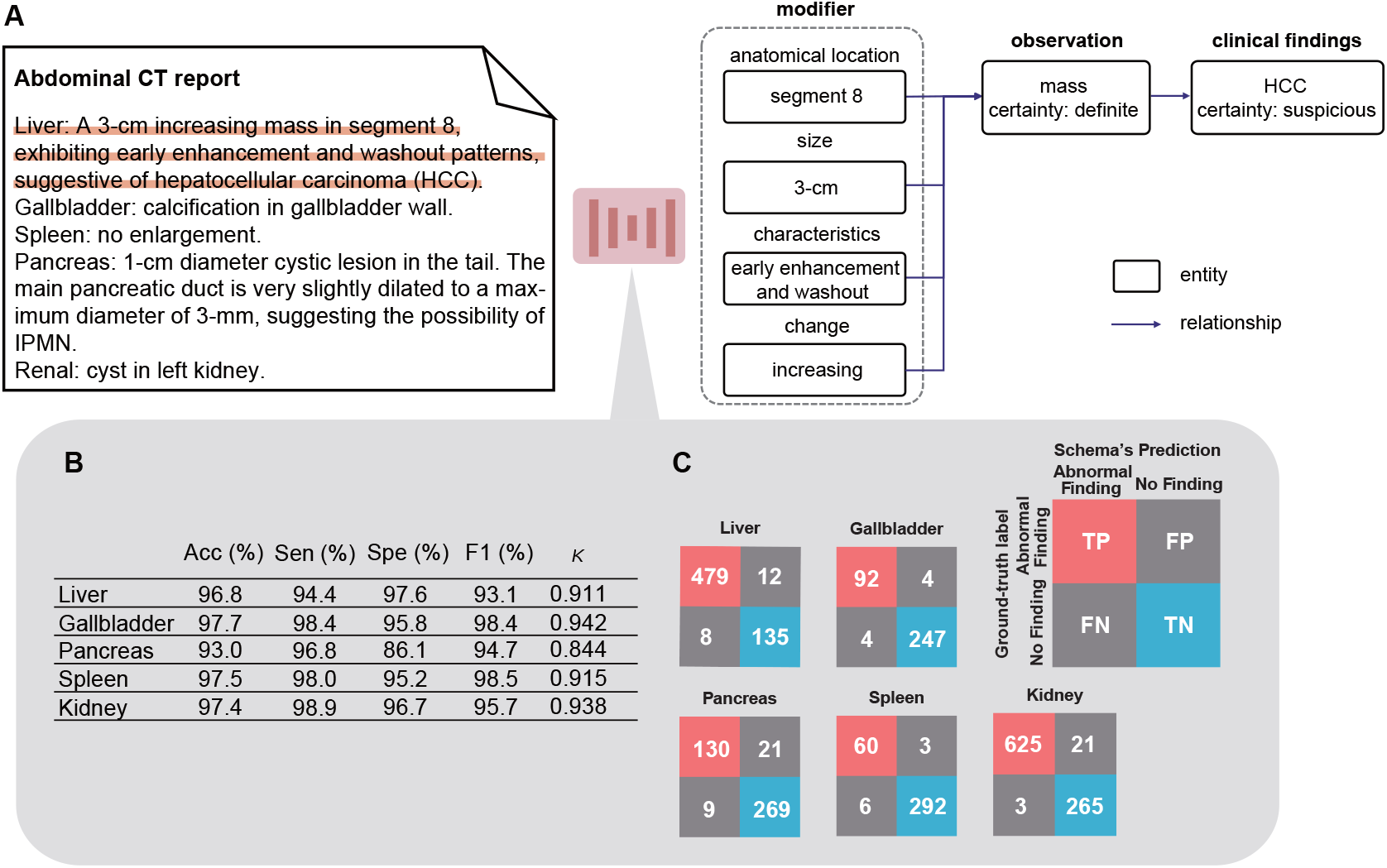
Representative example of the information-extraction schema applied to a free-text radiology report and illustration of the schema’s performance. (A) Our schema extracts imaging findings, associated information, and suspected diseases from radiology reports. (B) Disease extraction performance per organ by the schema is evaluated using accuracy (Acc), sensitivity (Sen), specificity (Spe), F1 score (F1), kappa coefficient, and confusion matrix values. (C) These confusion matrices cross-tabulate the true labels, manually labeled by radiologists, against predictions from the information extraction schema. True positives (TP) and true negatives (TN) represent accurately predicted cases, while false positives (FP) and false negatives (FN) highlight misclassifications.

### Anomaly detection performance of our model

Our anomaly detection model was trained using images from 66,684 exams (39,255 patients) across three institutions (Figure 1). This training cohort included 252,762 instances across all organs—48,848 liver instances (13,092 normal, 35,756 abnormal; 30,897 patients), 29,942 gallbladder instances (22,127 normal, 7,815 abnormal; 19,372 patients), 37,040 pancreas instances (28,116 normal, 8,924 abnormal; 22,605 patients), 32,465 spleen instances (28,069 normal, 4,396 abnormal; 19,960 patients), and 86,864 kidney instances (23,546 normal, 63,318 abnormal; 38.822 patients). We validated the model’s performance on an internal test cohort of 300 exams (295 individuals) across three institutions as well as an external test cohort of 600 exams (596 patients) from six hospitals. Details of patient characteristics, abnormal label ratio, and CT vendor information are listed in Supplementary Tables 2–5.

The AUCs for the internal and external test cohorts are shown in Figure 5. The average AUC for anomaly detection across organs in the external test cohort was 0·886: liver, 0·903 [95%CI: 0·877–0·929], gallbladder 0·898 [95%CI: 0·859–0·934], pancreas 0·838 [95%CI: 0·795–0·882], spleen 0·894 [95%CI: 0·845–0·938], and kidney, 0·898 [95%CI: 0·870–0·923]. The liver, gallbladder, and kidney had higher AUCs in the external cohort than in the internal cohort, with only a slight difference in the average AUC values between the internal (0·881) and external (0·886) cohorts. The accuracy, sensitivity, specificity, F1 score for the internal and external test cohorts are shown in Table 1, and the precision–recall AUCs are shown in Supplementary Figure 2.

**Table 1.** Anomaly detection performance metrics of the model for internal and external test cohorts.

|  |  | Accuracy, % | Sensitivity,<br>% | Specificity,<br>% | F1 score, % |
| --- | --- | --- | --- | --- | --- |
| Liver | External | 84.8<br>(82.0–87.5) | 88.9<br>(85.7–91.9) | 75.8<br>(70.0–81.9) | 89.0<br>(86.8–91.1) |
|  | Internal | 81.6<br>(77.3–85.6) | 92.6<br>(88.5–95.9) | 63.1<br>(54.1–71.8) | 86.4<br>(82.5–89.5) |
| Gallbladder | External | 86.6<br>(83.4–89.5) | 79.0<br>(71.8–85.8) | 89.0<br>(85.7–92.0) | 73.7<br>(67.1–79.4) |
|  | Internal | 83.5<br>(79.5–87.8) | 81.2<br>(71.7–89.9) | 84.3<br>(79.4–89.4) | 72.7<br>(64.9–79.8) |
| Pancreas | External | 81.7<br>(78.7–84.6) | 71.6<br>(63.7–78.6) | 84.6<br>(81.4–87.8) | 63.8<br>(56.9–69.7) |
|  | Internal | 79.6<br>(74.8–84.4) | 81.8<br>(70.8–91.8) | 79.1<br>(73.8–84.0) | 60<br>(49.6–68.9) |
| Spleen | External | 75.2<br>(71.6–78.6) | 85.5<br>(75.9–94.2) | 74.1<br>(70.1–77.7) | 39.2<br>(31.3–47.1) |
|  | Internal | 73.6<br>(68.5–78.8) | 96.8<br>(89.3–1) | 70.9<br>(65.3–76.3) | 43.8<br>(33.0–53.8) |
| Kidney | External | 76.8<br>(73.6–80.1) | 68.7<br>(64.1–73.2) | 91.9<br>(88.4–95.5) | 79.4<br>(76.1–82.7) |
|  | Internal | 76.5<br>(71.8–81.2) | 76.4<br>(70.6–82.0) | 76.8<br>(67.8–84.8) | 81.6<br>(77.4–85.3) |

Data are % (95% confidence interval).

**Figure 5.**
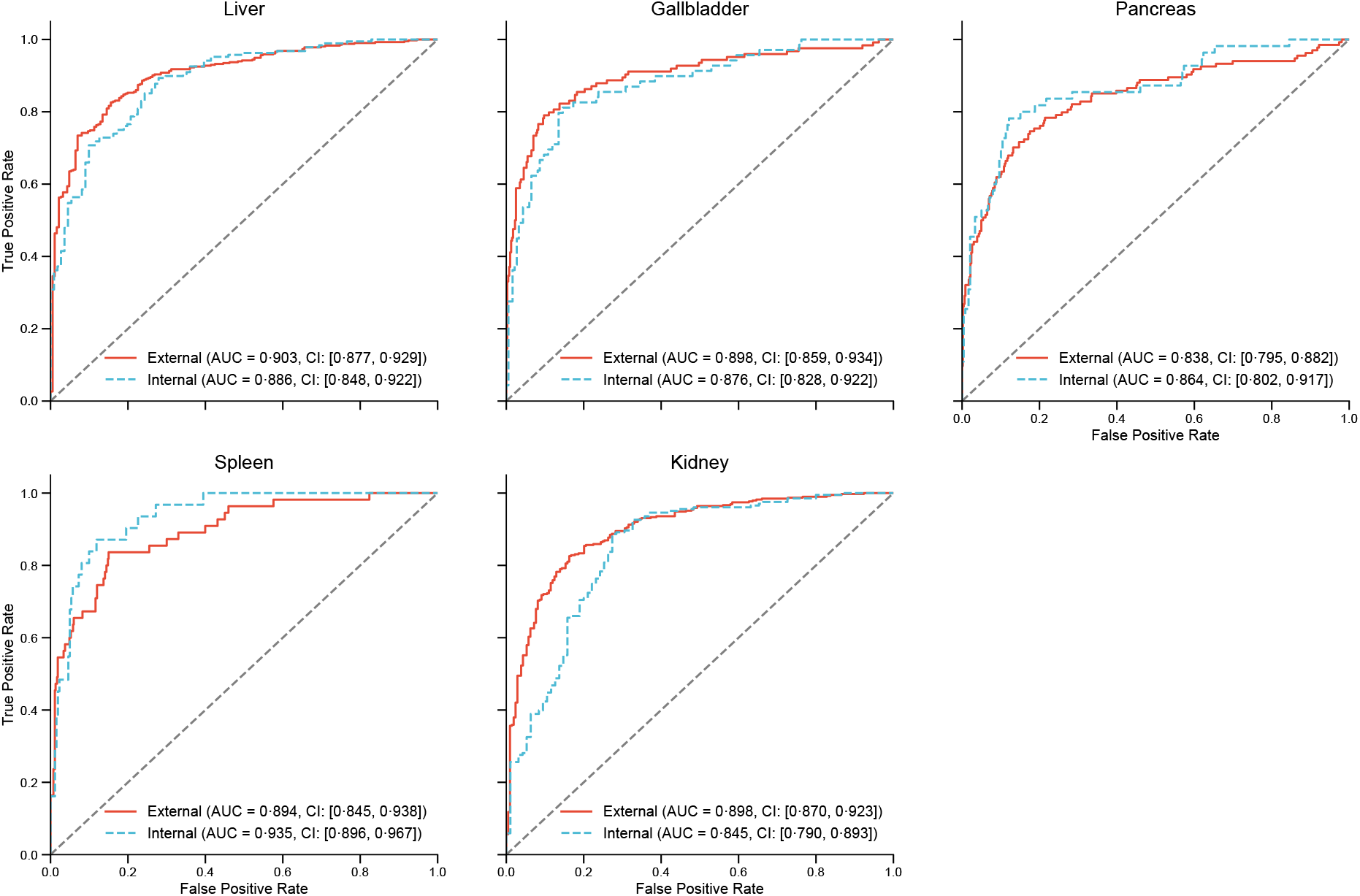
Receiver operating characteristic curves for the anomaly detection models for each abdominal organ Receiver operating characteristic curves for each organ in the internal and external test cohorts. The curve presents the true positive rate (sensitivity) and false positive rate (1 - specificity) across different cutoffs. The values in each graph represent the areas under the receiver operating characteristic curves (AUCs) and their 95% confidence intervals (CIs) for each cohort.

### Performance changes in different ratios of training data

A major advantage of our pipeline was its ability to use a large amount of data, obtained using an information extraction schema, without requiring manual annotations, for training. We then evaluated the anomaly detection performance using varying amounts of training data. We randomly selected 300, 1,000, 2,000, 6,000, and 12,000 exams for training across five organs and compared the AUCs obtained. Additionally, we trained models with ground-truth labels provided by radiologists for 300 exams. The results of the ROC curves and AUC for the external test data are shown in Figure 6. The models trained using 300 auto-labeled data points outperformed those trained using 300 expert-labeled data points. However, training models based on a larger dataset (≥ 2000) with auto-extracted labels significantly outperformed models trained with human labels, despite inaccuracies in the training labels.

**Figure 6.**
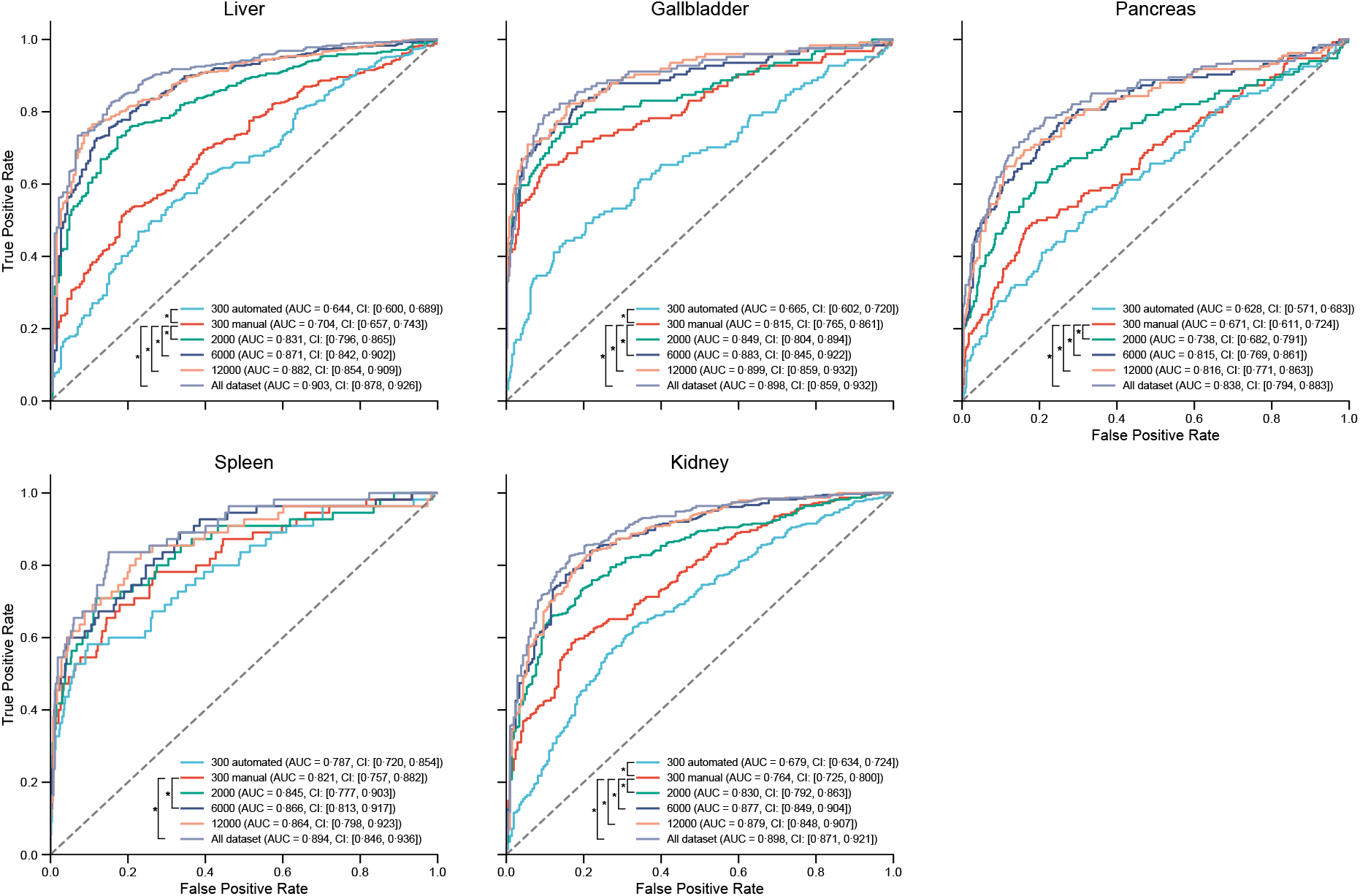
Impact of training data sizes on receiver operating characteristic curves. The receiver operating characteristic curve for each organ in the external test cohort illustrates the impact of varying training data sizes on the anomaly detection performance. The curve presents the true positive rate (sensitivity) and false positive rate (1 - specificity) across different cutoffs. The values in each graph represent the areas under the receiver operating characteristic curves (AUCs) and their 95% confidence intervals (CIs).

## Discussion

Here, we developed a deep-learning-based pipeline to detect clinical abnormalities in five organs to aid clinicians in diagnosis. A multi-organ segmentation model processed 3D CT images to extract organ-specific regions, and an information extraction schema was used to analyze the accompanying radiology reports to identify disease information. We then trained the anomaly detection models using organ-cropped images as the input and the presence of a disease as the training label. Our pipeline achieved an AUC of 0·886 against expert-annotated ground-truth labels in an external test cohort derived from six institutions. To the best of our knowledge, no previous study has extensively used free-text radiology reports as training labels for CT image classification, and our study involved the largest data set and number of anatomical sites analyzed in this way to date.

Previous studies using reports as labels have primarily focused on chest X-rays.^10–12^ Although a large amount of data for such 2D images is publicly available, these methods cannot be directly applied to 3D CT images because of differences in size and model structure. Only two studies to date have used reports as labels for CT images: one focused on the head region, with images collected from two institutions,^13^ while the other targeted two abdominal regions (liver/gallbladder and kidney), using a total of 9,153 images from a single institution, which yielded an anomaly-detection performance of at least 7% lower than that of our pipeline.^24^

Abnormality detection in abdominal CT images is challenging, because these scans provide high-resolution images of large areas including multiple organs. These images cannot be directly used in AI due to their size. Reducing their size compromises the details, while dividing them into small patches can prevent the model from recognizing structural abnormalities across all organs. Radiology reports detailing multiple organs reflect this complexity. To address these challenges, our pipeline employed segmentation models and an information extraction schema to identify and analyze these critical but small regions accurately across the organ spectrum. With the increasing availability of various open-source datasets for organ segmentation,^25^ applying our method to other organs might enable more extensive anomaly detection.

We demonstrated that the models trained on a large amount of data labeled by a language model outperformed those trained on smaller datasets labeled by experts, as shown in Figure 6. Additionally, compared with a previous study that used reports as training labels,^24^ our pipeline showed at least a 7% improvement in the AUC for organ-specific abnormality detection. This improvement may be due to the accuracy of the information extraction schema. A previous study on positron-emission tomography/CT applied information extraction algorithms to create labeled datasets from radiology reports to train an anomaly-detection model,^26^ and achieved a maximum F1 score of 0·888 in label creation. Our schema yielded higher F1 scores across all organs, suggesting that the automatic extraction of information from reports contributed to improved abnormality detection.

Extracting information from free-text radiology reports by employing deep-learning models, such as BERT,^10,11^ has proven superior to traditional rule-based methods.^7,27^ For reports with limited content, such as those for chest X-rays, direct input of the entire text into the model is sufficient for image-level classification.^10–12^ However, a different approach is required when identifying abnormalities in multiple organs, as in our case. In our approach, we used a language model to extract organ-specific abnormalities to create organ-specific training datasets and to extract information about the characteristics and longitudinal changes in abnormalities, which could facilitate improved accuracy in future. Additionally, our schema was language-independent, enabling its application to reports in various languages. With the emergence of large language models and the growing interest in leveraging existing medical data, our method could be clinically applicable.

Generalizability is a major challenge in deep-learning of medical images. Due to patient privacy and copyright concerns, data sharing is restricted, and we must often rely on datasets from a limited number of institutions. This scarcity of diverse datasets negatively affects model training and validation, leading to overfitting and good performance of models only on familiar images.^28^ While most available image data with accompanying radiological reports pertain to chest X-rays,^7,29^ no publicly available datasets of abdominal CT images with accompanying reports exist. Testing datasets from various institutions helps to avoid overfitting and ensures robustness and generalizability. We collected data from three institutions for our internal cohort and from six institutions for our external cohort, demonstrating the stable anomaly detection capabilities of our model across different imaging environments.

Our pipeline offers several advantages in clinical settings. First, our end-to-end pipeline enables the direct input of clinically acquired images without any pre-processing or annotation, which significantly reduces the development costs associated with clinical implementation. Moreover, the high AUC of our model addresses the challenge of increased imaging exams by reducing radiologist reading times. For example, AI for detecting breast cancer in digital breast tomosynthesis^30^ and lung nodule detection in chest radiographs^5^, with AUCs of 0·840 and > 0·9, respectively, significantly shortened reading times. Such diagnostic support systems provide strategic solutions to meet the growing demand for medical imaging.

Our study has several limitations. Retrospective data collection from multiple institutions suggested the need for further validation using prospectively collected test datasets across different disease prevalence rates. The training and evaluation of our model relied on diagnoses made by experienced radiologists, rather than final pathological diagnoses, which could misrepresent the actual abnormalities. A selection bias might also have been present, influenced by the protocol names and results of our information extraction schema. Additionally, exclusion of patients’ historical imaging data disallowed leveraging of temporal changes for anomaly detection, potentially limiting the model’s ability to reflect real-world clinical scenarios.

In conclusion, we developed a deep-learning-based pipeline encompassing labeled dataset creation, model training, and anomaly detection using CT images and associated free-text radiology reports. The learning process was streamlined by eliminating the need for manual annotations. Our pipeline was trained on a diverse dataset containing 252,762 imaging instances for five organs, which were collected from multiple institutions, and demonstrated high anomaly-detection capabilities for every organ examined. Our approach is broadly applicable across different anatomical sites and diseases, heralding significant advances in computer-aided diagnosis.

## Data Availability

All codes associated with pipeline development are shared on GitHub (https://github.com/jun-sato/sato_j-mid_ad). The pretrained models for both the multi-organ segmentation module and the anomaly detection module are available from JS. CT images are not available for sharing at this time, to protect the privacy of the participants.

## Acknowledgement

The manuscript was grammatically reviewed by ChatGPT.

## Contributors

All authors contributed to the conception and design of this study. JS and YS collected, verified, and analyzed the data, and provided access to the raw data. The ground-truth labels of the test data were created by JS, TW, DN, MT, and YH. KS and TT created the information extraction schema. JS wrote the first draft of the manuscript, which was then edited by YS, KS, and SK. All the processes were supervised by YS, TT, MH, SK, and NT. All authors had access to all data, and read and approved the final manuscript. SK was responsible for the decision to submit the manuscript for publication.

## Data Sharing

All codes associated with pipeline development are shared on GitHub (https://github.com/jun-sato/sato_j-mid_ad). The pretrained models for both the multiorgan segmentation module and the anomaly detection module are available from JS. CT images are not available for sharing at this time, to protect the privacy of the participants.

