## Supplementary Material for "Annotation-free multi-organ anomaly detection in abdominal CT using free-text radiology reports: A multi-center retrospective study"

### Contents of Supplementary Information

Supplementary Figure 1 – page 2

Supplementary Figure 2 – page 3

Supplementary Table 1 – page 4

Supplementary Table 2 – page 5,6

Supplementary Table 3 – page 7,8

Supplementary Table 4 – page 9

Supplementary Table 5 – page 10,11

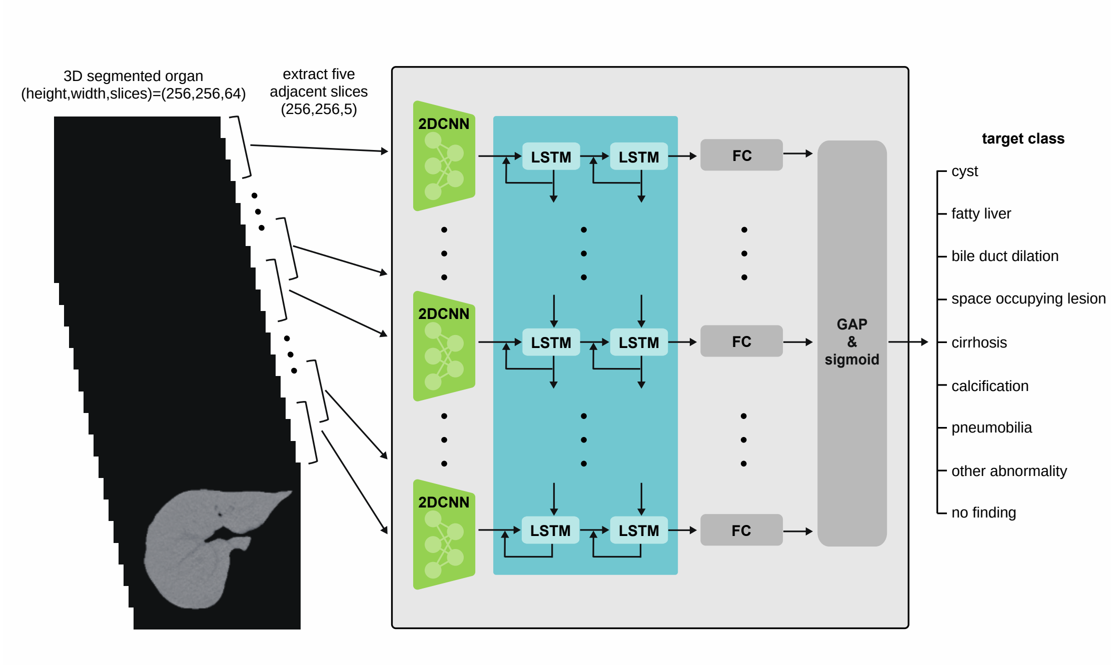


**Supplementary Figure 1: Outline of the anomaly detection module.**

The figure shows a schematic diagram of our deep learning model for detecting abnormal findings. Our model employs a multiple instance learning model to integrate different slice images. Five adjacent slices of segmented organ images are sequentially extracted with an overlap of two slices and input into a 2D convolutional neural network (CNN). Outputs from the CNN are fed into a Long Short-Term Memory (LSTM) network to share information between slices. Fully connected layers (FC) then act as a classifier. Information across slices is aggregated using global average pooling (GAP), followed by a sigmoid activation function and a cross-entropy loss function. During training, the model is trained as a multi-class classifier including no findings and multiple abnormal findings. During inference, it simplifies to binary classification, detecting only the presence or absence of anomalies.

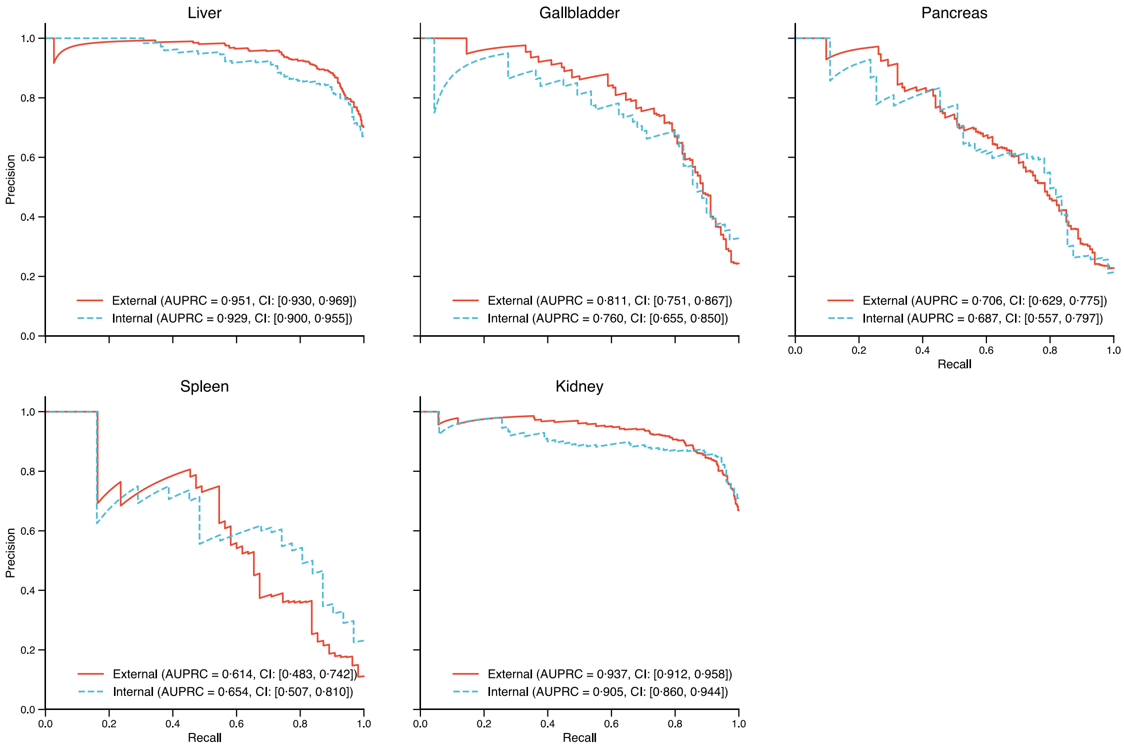


**Supplementary Figure 2: Precision-recall curves for the anomaly detection models**

The precision-recall curve for each organ in the internal and external test cohort. The curve presents the precision and recall across different cutoffs. The values on each graph are the areas under the precision-recall curve (AUPRCs) and their 95% confidence intervals for each cohort.

**Supplementary Table 1: Performance of multi-organ segmentation model**

| | Liver | Spleen | Pancreas | Gallbladder | Left kidney | Right kidney |
|---|---|---|---|---|---|---|
| **DSC** | 0·981 (0·974–0·986) | 0·979 (0·972–0·985) | 0·911 (0·882–0·934) | 0·937 (0·913–0·957) | 0·973 (0·963–0·980) | 0·972 (0·965–0·979) |
| **NSD** | 0·871 (0·820–0·937) | 0·934 (0·902–0·965) | 0·77 (0·706–0·831) | 0·865 (0·784–0·935) | 0·918 (0·887–0·952) | 0·923 (0·893–0·950) |

Data are median (IQR). DSC=dice similarity coefficient. NSD=normalized surface dice.

**Supplementary Table 2: Images per vendor in the internal training cohort**

| | Manufacturer | Model Name | Keio | Okayama | Tokyo |
|---|---|---|---|---|---|
| **Liver** | Canon Medical Systems | Aquilion ONE | 273 | 2702 | 1300 |
| | | Aquilion Precision | | 2527 | 2336 |
| | | Aquilion PRIME | | | 8905 |
| | | Aquilion Prime SP | | | 1858 |
| | GE Healthcare | Discovery CT750 HD | 4256 | 2678 | 5318 |
| | | Revolution CT | 3190 | | 8164 |
| | | Revolution EVO | 1063 | | |
| | | BrightSpeed | 601 | | |
| | Siemens | SOMATOM Definition | | 2568 | |
| | | SOMATOM go.Top | | 1030 | |
| | | NAEOTOM Alpha | | 79 | |
| **Gallbladder** | Canon Medical Systems | Aquilion ONE | 54 | 1064 | 1094 |
| | | Aquilion Precision | | 902 | 1986 |
| | | Aquilion PRIME | | | 7576 |
| | | Aquilion Prime SP | | | 1524 |
| | GE Healthcare | Discovery CT750 HD | 646 | 1128 | 4739 |
| | | Revolution CT | 540 | | 6957 |
| | | Revolution EVO | 264 | | |
| | | BrightSpeed | 37 | | |
| | Siemens | SOMATOM Definition | | 978 | |
| | | SOMATOM go.Top | | 412 | |
| | | NAEOTOM Alpha | | 41 | |
| **Pancreas** | Canon Medical Systems | Aquilion ONE | 105 | 1235 | 1399 |
| | | Aquilion Precision | | 1171 | 2508 |
| | | Aquilion PRIME | | | 9290 |
| | | Aquilion Prime SP | | | 1867 |
| | GE Healthcare | Discovery CT750 HD | 1045 | 1315 | 5942 |
| | | Revolution CT | 783 | | 8418 |
| | | Revolution EVO | 297 | | |
| | | BrightSpeed | 39 | | |
| | Siemens | SOMATOM Definition | | 1166 | |
| | | SOMATOM go.Top | | 414 | |
| | | NAEOTOM Alpha | | 46 | |
| **Spleen** | Canon Medical Systems | Aquilion ONE | 39 | 901 | 1326 |
| | | Aquilion Precision | | 817 | 2304 |
| | | Aquilion PRIME | | | 8645 |
| | | Aquilion Prime SP | | | 1756 |
| | GE Healthcare | Discovery CT750 HD | 446 | 971 | 5569 |
| | | Revolution CT | 352 | | 7904 |
| | | Revolution EVO | 199 | | |
| | | BrightSpeed | 22 | | |
| | Siemens | SOMATOM Definition | | 865 | |
| | | SOMATOM go.Top | | 315 | |
| | | NAEOTOM Alpha | | 34 | |

| | | | | | |
|---|---|---|---|---|---|
| **Kidney** | Canon Medical Systems | Aquilion ONE | 411 | 4516 | 2407 |
| | | Aquilion Precision | | 4261 | 4638 |
| | | Aquilion PRIME | | | 17102 |
| | | Aquilion Prime SP | | | 3225 |
| | GE Healthcare | Discovery CT750 HD | 6451 | 4798 | 10437 |
| | | Revolution CT | 4997 | | 15018 |
| | | Revolution EVO | 1788 | | |
| | | BrightSpeed | 441 | | |
| | Siemens | SOMATOM Definition | | 4488 | |
| | | SOMATOM go.Top | | 1734 | |
| | | NAEOTOM Alpha | | 148 | |
| | | Unknown | | 4 | |

Data are n.

#### Supplementary Table 3: Images per vendor in the internal and external test cohort

| | Manufacturer | Model Name | Tokyo | Keio | Okayama | Ehime | Juntendo | Kyoto | Kyushu | Osaka | Tokushima |
|---|---|---|---|---|---|---|---|---|---|---|---|
| **Liver** | Canon Medical Systems | Aquilion ONE | 15 | 3 | 30 | 29 | 89 | 41 | 58 | 39 | 100 |
| | | Aquilion Precision | | | 16 | | | | 19 | 29 | |
| | | Aquilion PRIME | 10 | | | | | 59 | 1 | | |
| | | Aquilion Prime SP | 34 | | | | | | | | |
| | GE Healthcare | Discovery CT750 HD | 13 | 44 | 24 | | | | | | |
| | | Revolution CT | 28 | 36 | | | | | | 32 | |
| | | Revolution EVO | | 12 | | | | | | | |
| | | BrightSpeed | | 5 | | | | | | | |
| | Siemens | SOMATOM go.Top | | | 9 | | | | | | |
| | | SOMATOM Force | | | | 45 | | | | | |
| | | NAEOTOM Alpha | | | 21 | | | | | | |
| | Philips | iCT 256 | | | | 26 | 11 | | | | |
| | | IQon - Spectral CT | | | | | | | 22 | | |
| **Gallbladder** | Canon Medical Systems | Aquilion ONE | 13 | 2 | 25 | 23 | 85 | 40 | 48 | 33 | 92 |
| | | Aquilion Precision | | | 14 | | | | 15 | 24 | |
| | | Aquilion PRIME | 9 | | | | | 50 | | | |
| | | Aquilion Prime SP | 30 | | | | | | | | |
| | GE Healthcare | Discovery CT750 HD | 12 | 41 | 22 | | | | | | |
| | | Revolution CT | 27 | 30 | | | | | | 28 | |
| | | Revolution EVO | | 9 | | | | | | | |
| | | BrightSpeed | | 5 | | | | | | | |
| | Siemens | SOMATOM go.Top | | | 8 | | | | | | |
| | | SOMATOM Force | | | | 40 | | | | | |
| | | NAEOTOM Alpha | | | 20 | | | | | | |
| | Philips | iCT 256 | | | | 25 | 11 | | | | |
| | | IQon - Spectral CT | | | | | | | 19 | | |
| **Pancreas** | Canon Medical Systems | Aquilion ONE | 15 | 3 | 30 | 29 | 89 | 41 | 58 | 39 | 100 |
| | | Aquilion Precision | | | 15 | | | | 19 | 29 | |
| | | Aquilion PRIME | 10 | | | | | 59 | 1 | | |
| | | Aquilion Prime SP | 34 | | | | | | | | |
| | GE Healthcare | Discovery CT750 HD | 13 | 44 | 24 | | | | | | |
| | | Revolution CT | 28 | 36 | | | | | | 32 | |

| | | | | | | | | | | | |
|---|---|---|---|---|---|---|---|---|---|---|---|
| | | Revolution EVO | | 12 | | | | | | | |
| | | BrightSpeed | | 5 | | | | | | | |
| | Siemens | SOMATOM go.Top | | | 9 | | | | | | |
| | | SOMATOM Force | | | | 45 | | | | | |
| | | NAEOTOM Alpha | | | 21 | | | | | | |
| | Philips | iCT 256 | | | | 26 | 11 | | | | |
| | | IQon - Spectral CT | | | | | | | 22 | | |
| **Spleen** | Canon Medical Systems | Aquilion ONE | 15 | 3 | 30 | 29 | 89 | 40 | 58 | 35 | 100 |
| | | Aquilion Precision | | | 15 | | | | 19 | 29 | |
| | | Aquilion PRIME | 10 | | | | | 58 | | | |
| | | Aquilion Prime SP | 34 | | | | | | | | |
| | GE Healthcare | Discovery CT750 HD | 13 | 42 | 24 | | | | | | |
| | | Revolution CT | 28 | 35 | | | | | | 31 | |
| | | Revolution EVO | | 12 | | | | | | | |
| | | BrightSpeed | | 5 | | | | | | | |
| | Siemens | SOMATOM go.Top | | | 9 | | | | | | |
| | | SOMATOM Force | | | | 45 | | | | | |
| | | NAEOTOM Alpha | | | 21 | | | | | | |
| | Philips | iCT 256 | | | | 26 | 11 | | | | |
| | | IQon - Spectral CT | | | | | | | 22 | | |
| **Kidney** | Canon Medical Systems | Aquilion ONE | 30 | 6 | 60 | 56 | 176 | 81 | 113 | 76 | 198 |
| | | Aquilion Precision | | | 31 | | | | 38 | 57 | |
| | | Aquilion PRIME | 20 | | | | | 117 | 2 | | |
| | | Aquilion Prime SP | 67 | | | | | | | | |
| | GE Healthcare | Discovery CT750 HD | 26 | 86 | 47 | | | | | | |
| | | Revolution CT | 55 | 72 | | | | | | 62 | |
| | | Revolution EVO | | 24 | | | | | | | |
| | | BrightSpeed | | 10 | | | | | | | |
| | Siemens | SOMATOM go.Top | | | 18 | | | | | | |
| | | SOMATOM Force | | | | 88 | | | | | |
| | | NAEOTOM Alpha | | | 42 | | | | | | |
| | Philips | iCT 256 | | | | 49 | 21 | | | | |
| | | IQon - Spectral CT | | | | | | | 44 | | |

Data are n.

**Supplementary Table 4: Patient characteristics in the internal training cohort**

| | | | Liver | Gallbladder | Pancreas | Spleen | Kidney |
|---|---|---|---|---|---|---|---|
| **Tokyo** | Exams | | 27,881 | 23,876 | 29,424 | 27,504 | 52,827 |
| | Patients | | 17,172 | 15,135 | 17,602 | 16,645 | 17,420 |
| | Abnormal label | | 17177 (61.6) | 4499 (18.8) | 4634 (15.7) | 2429 (8.8) | 21579 (40.8) |
| | Sex | Male | 15841 (56.8) | 13273 (55.6) | 16725 (56.8) | 15582 (56.7) | 29869 (56.5) |
| | | Female | 12040 (43.2) | 10603 (44.4) | 12699 (43.2) | 11922 (43.3) | 22958 (43.5) |
| | Age, years | | 68 (55–77) | 67 (54–76) | 68 (55–76) | 68 (55–76) | 68 (54–76) |
| **Keio** | Exams | | 9,383 | 1,541 | 2,269 | 1,058 | 14,088 |
| | Patients | | 6,347 | 1,157 | 1,498 | 745 | 6,016 |
| | Abnormal label | | 9178 (97.8) | 1359 (88.2) | 2014 (88.8) | 917 (86.7) | 13782 (97.8) |
| | Sex | Male | 4899 (52.2) | 895 (58.1) | 1330 (58.6) | 617 (58.3) | 8759 (62.2) |
| | | Female | 4484 (47.8) | 646 (41.9) | 939 (41.4) | 441 (41.7) | 5329 (37.8) |
| | Age, years | | 67 (56–76) | 70 (59–78) | 72 (63–79) | 64 (51–73) | 71 (61–78) |
| **Okayama** | Exams | | 11,584 | 4,525 | 5,347 | 3,903 | 19,949 |
| | Patients | | 7,378 | 3,080 | 3,505 | 2,570 | 7,232 |
| | Abnormal label | | 9401 (81.2) | 1957 (43.2) | 2276 (42.6) | 1050 (26.9) | 12717 (63.7) |
| | Sex | Male | 6268 (54.1) | 2477 (54.7) | 2980 (55.7) | 2172 (55.6) | 11488 (57.6) |
| | | Female | 5315 (45.9) | 2048 (45.3) | 2367 (44.3) | 1731 (44.4) | 8459 (42.4) |
| | | Unknown | 1 | | | | 2 |
| | Age, years | | 68 (56–75) | 67 (54–74) | 69 (58–76) | 67 (53–74) | 69 (58–76) |
| **Overall** | Exams | | 48,848 | 29,942 | 37,040 | 32,465 | 86,864 |
| | Patients | | 30,897 | 19,372 | 22,605 | 19,960 | 30,668 |
| | Abnormal label | | 35756 (73.2) | 7815 (26.1) | 8924 (24.1) | 4396 (13.5) | 63318 (60.6) |
| | Sex | Male | 27008 (55.3) | 16645 (55.6) | 21035 (56.8) | 18371 (56.6) | 50116 (57.7) |
| | | Female | 21839 (44.7) | 13297 (44.4) | 16005 (43.2) | 14094 (43.4) | 36746 (42.3) |
| | | Unknown | 1 | | | | 2 |
| | Age, years | | 68 (56–76) | 67 (54–76) | 69 (56–77) | 68 (54–76) | 69 (57–76) |

Data are n, n (%), or median (IQR).

**Supplementary Table 5: Patient characteristics in the internal and external test cohort**

| | | | | Liver | Gallbladder | Pancreas | Spleen | Kidney |
|---|---|---|---|---|---|---|---|---|
| **Internal** | **Tokyo** | Exams | | 100 | 91 | 100 | 100 | 198 |
| | | Patients | | 98 | 90 | 98 | 98 | 98 |
| | | Abnormal label | | 52 (52.0) | 22 (24.2) | 88 (88.0) | 8 (8.0) | 89 (44.9) |
| | | Sex | Male | 63 (63.0) | 58 (63.7) | 63 (63.0) | 63 (63.0) | 124 (62.6) |
| | | | Female | 37 (37.0) | 33 (36.3) | 37 (37.0) | 37 (37.0) | 74 (37.4) |
| | | Age, years | | 69(57–76) | 68(54–76) | 69(57–76) | 69(57–76) | 69(56–76) |
| | **Keio** | Exams | | 100 | 87 | 100 | 97 | 198 |
| | | Patients | | 97 | 85 | 97 | 95 | 97 |
| | | Abnormal label | | 65 (65.0) | 28 (32.2) | 25 (25.0) | 9 (9.3) | 137 (69.2) |
| | | Sex | Male | 55 (55.0) | 45 (51.7) | 55 (55.0) | 52 (53.6) | 108 (54.5) |
| | | | Female | 45 (45.0) | 42 (48.3) | 45 (45.0) | 45 (46.4) | 90 (45.5) |
| | | Age, years | | 68(57–74) | 64(56–74) | 68(57–74) | 67(56–74) | 69(57–74) |
| | **Okayama** | Exams | | 100 | 89 | 99 | 99 | 198 |
| | | Patients | | 100 | 89 | 99 | 99 | 100 |
| | | Abnormal label | | 72 (72.0) | 22 (24.7) | 18 (18.2) | 14 (14.1) | 104 (52.5) |
| | | Sex | Male | 52 (52.0) | 48 (53.9) | 52 (52.5) | 52 (52.5) | 102 (51.5) |
| | | | Female | 48 (48.0) | 41 (46.1) | 47 (47.5) | 47 (47.5) | 96 (48.5) |
| | | Age, years | | 71(53–77) | 70(53–77) | 71(53–77) | 71(53–77) | 71(53–77) |
| **External** | **Ehime** | Exams | | 100 | 88 | 100 | 100 | 193 |
| | | Patients | | 100 | 88 | 100 | 100 | 100 |
| | | Abnormal label | | 73 (73.0) | 28 (31.8) | 23 (23.0) | 8 (8.0) | 103 (53.4) |
| | | Sex | Male | 53 (53.0) | 45 (51.1) | 53 (53.0) | 53 (53.0) | 103 (53.4) |
| | | | Female | 47 (47.0) | 43 (48.9) | 47 (47.0) | 47 (47.0) | 90 (46.6) |
| | | Age, years | | NA | NA | NA | NA | NA |
| | **Juntendo** | Exams | | 100 | 96 | 100 | 100 | 197 |
| | | Patients | | 99 | 95 | 99 | 99 | 99 |
| | | Abnormal label | | 64 (64.0) | 11 (11.5) | 10 (10.0) | 5 (5.0) | 98 (49.7) |
| | | Sex | Male | 62 (62.0) | 60 (62.5) | 62 (62.0) | 62 (62.0) | 122 (61.9) |
| | | | Female | 38 (38.0) | 36 (37.5) | 38 (38.0) | 38 (38.0) | 75 (38.1) |
| | | Age, years | | 63(56–74) | 63(56–72) | 63(56–74) | 63(56–74) | 63(56–74) |

| | | | | | | | |
|---|---|---|---|---|---|---|---|
| **Kyoto** | Exams | | 100 | 90 | 100 | 98 | 198 |
| | Patients | | 98 | 88 | 98 | 96 | 98 |
| | Abnormal label | | 78 (78.0) | 27 (30.0) | 19 (19.0) | 5 (5.1) | 115 (58.1) |
| | Sex | Male | 56 (56.0) | 51 (56.7) | 56 (56.0) | 54 (55.1) | 111 (56.1) |
| | | Female | 44 (44.0) | 39 (43.3) | 44 (44.0) | 44 (44.9) | 87 (43.9) |
| | Age, years | | 72(56–78) | 73(57–78) | 72(56–78) | 73(56–78) | 71(56–78) |
| **Kyushu** | Exams | | 100 | 82 | 100 | 99 | 197 |
| | Patients | | 100 | 82 | 100 | 99 | 100 |
| | Abnormal label | | 61 (61.0) | 13 (15.9) | 26 (26.0) | 12 (12.1) | 99 (50.3) |
| | Sex | Male | 58 (58.0) | 49 (59.8) | 58 (58.0) | 58 (58.6) | 116 (58.9) |
| | | Female | 42 (42.0) | 33 40.2) | 42 (42.0) | 41 (41.4) | 81 (41.1) |
| | Age, years | | 66(52–71 | 65(49–71) | 66(52–71) | 66(52–71) | 65(50–71) |
| **Osaka** | Exams | | 100 | 85 | 100 | 95 | 195 |
| | Patients | | 99 | 84 | 99 | 94 | 99 |
| | Abnormal label | | 73 (73.0) | 23 (27.1) | 33 (33.0) | 13 (13.7) | 108 (55.4) |
| | Sex | Male | 64 (64.0) | 55 (64.7) | 64 (64.0) | 61 (64.2) | 124 (63.6) |
| | | Female | 36 (36.0) | 30 (35.3) | 36 (36.0) | 34 (35.8) | 71 (36.4) |
| | Age, years | | 68(59–77) | 66(59–76) | 68(59–77) | 77(59–77) | 68(59–77) |
| **Tokushima** | Exams | | 100 | 92 | 100 | 100 | 198 |
| | Patients | | 100 | 92 | 100 | 100 | 100 |
| | Abnormal label | | 63 (63.0) | 25 (27.2) | 25 (25.0) | 12 (12.0) | 124 (62.6) |
| | Sex | Male | 65 (65.0) | 59 (64.1) | 65 (65.0) | 65 (65.0) | 130 (65.7) |
| | | Female | 35 (35.0) | 33 (35.9) | 35 (35.0) | 35 (35.0) | 68 (34.3) |
| | Age, years | | 71(59–78) | 71(57–77) | 71(59–78) | 71(59–78) | 71(59–78) |
| **Overall exams** | Exams | | 900 | 800 | 899 | 888 | 1,772 |
| | Patients | | 891 | 793 | 890 | 880 | 891 |
| | Abnormal label | | 603 (67.0) | 199 (24.9) | 191 (21.2) | 86 (9.7) | 977 (55.1) |
| | Sex | Male | 524 (58.2) | 463 (57.9) | 523 (58.2) | 515 (58.0) | 1034 (58.4) |
| | | Female | 376 (41.8) | 337 (42.1) | 376 (41.8) | 373 (42.0) | 738 (41.6) |
| | Age, years | | 69(56–76) | 67(56–76) | 68(56–76) | 68(56–76) | 68(56–76) |

Data are n, n (%), or median (IQR).
